# A differential effect of visual cortex tDCS on reading of English and Chinese in patients with central vision loss

**DOI:** 10.1101/2022.07.24.22277956

**Authors:** A.E. Silva, A. Lyu, S.J. Leat, S. Khan, T. Labreche, J.C.H. Chan, Q. Li, G.C. Woo, S. Woo, A.M.Y Cheong, B. Thompson

## Abstract

We report that visual cortex a-tDCS influences reading performance in individuals with macular degeneration differently depending on the writing system employed. This finding will help to guide the international development of vision rehabilitation programs for macular degeneration that may utilize non-invasive brain stimulation.

---

Peripheral vision is susceptible to crowding, a difficulty in distinguishing neighbouring objects or features (Bouma, 1970). Crowding impairs face and object recognition and reading in individuals with central vision loss who are forced to rely on peripheral vision (Wallace et al., 2017). Anodal transcranial direct current stimulation (a-tDCS) of primary visual cortex reduces crowding in normal peripheral vision (Chen et al., 2021). Furthermore, a recent case series suggested that visual cortex a-tDCS reduces collinear inhibition, a phenomenon related to crowding, in patients with central vision loss due to macular degeneration (Raveendran et al., 2021). Building on this prior work, in this pre-registered (ClinicalTrials.gov ID: NCT04111068), within-subjects, randomized, double-blind, placebo-controlled study, we tested the hypothesis that reading would be improved during, immediately after, and 30 minutes after visual cortex a-tDCS in adults with central vision loss due to macular degeneration. We also assessed whether the effect of a-tDCS would vary between two writing systems, an alphabet system (English written in the Roman alphabet) and a logographic system (traditional Chinese characters). Within a single alphabetic word, crowding impairs discrimination of adjacent letters. Within a single logographic character, crowding impairs discrimination of internal structure. We hypothesised that a-tDCS would improve reading for both writing systems by reducing crowding.

Twenty-one individuals (8 females, age 72 ± 14 years) with macular degeneration participated. Using their better eye, 8 read English words and 13 read Chinese characters. Full methods are provided in the supplementary materials. All participants provided written informed consent, the study was reviewed and received ethics clearance through the University of Waterloo and The Hong Kong Polytechnic University Research Ethics Boards, and all procedures adhered to the tenets of the Declaration of Helsinki.

Participants performed a rapid serial visual presentation (RSVP) verbal reading task whereby each word or character in a sentence was presented sequentially at the same location (Harland et al., 1998). This method measures reading ability without the confounding effect of reading-related eye movements. Print size and exposure duration were individually selected to elicit 55% reading accuracy prior to the main experimental task.

In each test block (pre, 5 min post and 30 min post stimulation), participants read 15 sentences. Accuracy was measured as the total proportion of words (English) or characters (Chinese) read correctly. Participants completed both an active and a sham (placebo) a-tDCS session separated by at least 48 hours. The order of stimulation type was randomized. A-tDCS (2 mA, 20 min, 30 sec ramp up/down) was administered using a neuroConn DC-Stimulator plus (Waterloo) or Neuro Device Group S.A. nurostym tES (Hong Kong) and two 5 cm × 5 cm rubber electrodes inside saline soaked sponges. The anode was placed over Oz and the cathode was placed over a randomly selected cheek, both secured using head straps (Reinhart et al., 2016). Sham stimulation involved the 30 second ramps only.

Following a pre-specified analysis plan, the effect of brain stimulation was calculated by subtracting the pre-test reading accuracy from that of each post-test for each participant. A 2 (Stimulation type: Active, Sham) × 3 (Test: During, Post 5, Post 30) repeated-measures ANOVA with language included as a dichotomous covariate revealed a significant interaction between stimulation type and language, *F*(1,19) = 4.7, *p* = 0.043. Fig 1 illustrates that active stimulation increased reading accuracy relative to sham in participants reading English sentences but did not increase accuracy in participants reading Chinese characters. Separate analyses of each language group did not reveal any statistically significant effects, likely due to the limited sample size. Secondary outcome measures included crowded and uncrowded Landolt C visual acuity, as measured by the Freiberg Vision Test (FrACT) (Bach, 1996). A similar pattern of results was observed for crowded visual acuity (See Supplementary Results).

**Fig 1:**
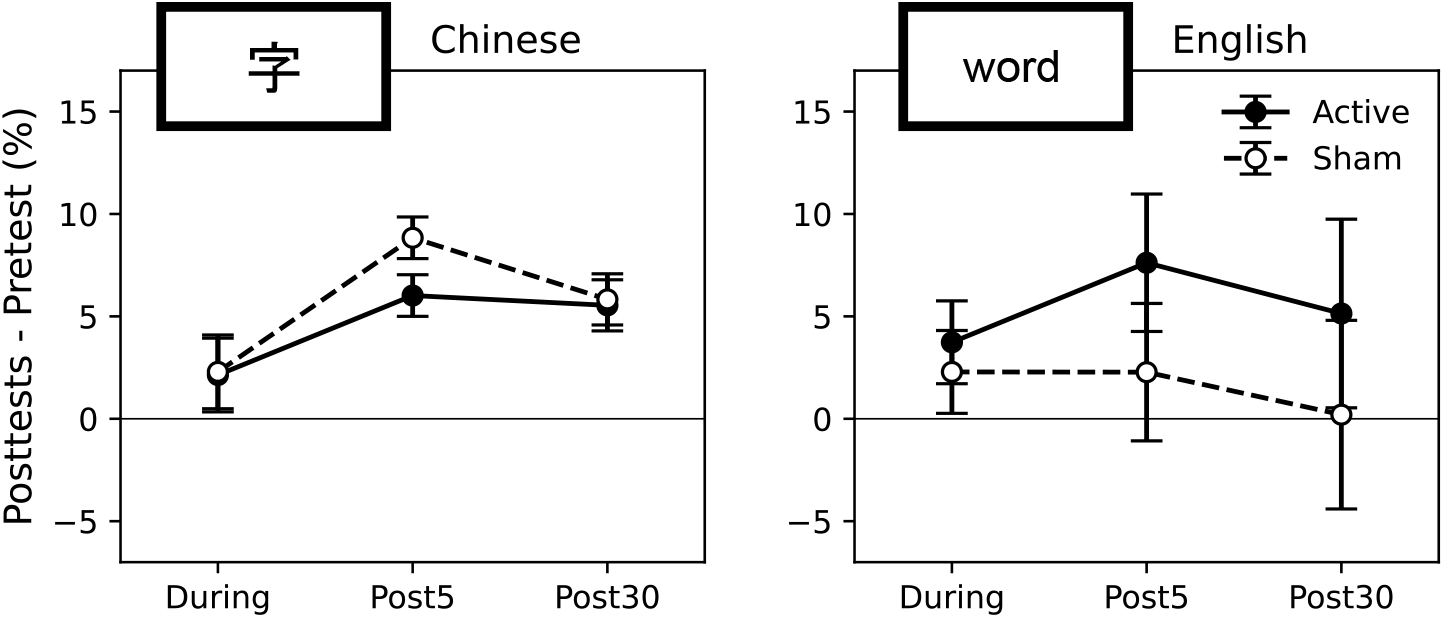
The effect of a-tDCS on reading accuracy for Chinese characters (left plot) and English words (right plot). Sentences were presented one word or character at a time (stimulus inserts). Participants were tested before, during, 5 minutes after, and 30 minutes after application of tDCS. The Y-axis denotes the accuracy difference between the pre-test and each of the three post-tests. The error bars are ±1 SEM.

These results demonstrate that visual cortex a-tDCS influences reading performance in individuals with macular degeneration differently depending on writing system. This observation will help to guide the international development of vision rehabilitation programs for macular degeneration that may utilize non-invasive brain stimulation. Importantly, our results relate to the acute effects of a-tDCS on reading performance that we assume to be mediated by an immediate reduction of between-letter crowding in English sentences. While no improvement was observed when reading individual Chinese characters, the effect of a-tDCS on between-character crowding within strings of characters remains an open question. There is strong evidence that non-invasive brain stimulation techniques, including a-tDCS, can enhance perceptual learning of visual tasks in individuals with normal or impaired vision (Contemori et al., 2019; Spiegel et al., 2013; Yang et al., 2022). Exploration of whether a-tDCS can increase the rate and magnitude of a reading-based perceptual learning task and whether any effects are modulated by writing system is warranted.

## Supporting information

supplementary matrials

## Data Availability

All data produced in the study are available upon reasonable request, subject to the study Steering Group's decision.

## Acknowledgments

This research is supported by CFI (34095), NSERC (RPIN-05394 and RGPAS-477166), and the Velux Stiftung Foundation (1188). This research is also supported by InnoHK and the Hong Kong SAR government. AES is supported by a Transformative Quantum Technologies postdoctoral fellowship.

