## supplementary matrials for "A differential effect of visual cortex tDCS on reading of English and Chinese in patients with central vision loss"

### Supplementary Methods

#### Participants

Inclusion criteria were central vision loss due to age-related macular degeneration (AMD) or juvenile macular degeneration (JMD), best-corrected distance visual acuity (BCVA) between 0.18 and 1.20 logMAR and 0.4M at 40 cm in the better eye, and participant-reported stable vision for the last 3 months.

Exclusion criteria included a diagnosis of dementia, not being fluent in English (Waterloo) or Chinese character reading (Hong Kong), any ocular surgery (including anti-VEGF injections) within the duration of the study, ocular pathology other than AMD or JMD that significantly reduced central vision, a severe hearing impairment, and any contraindications for tDCS.

At the University of Waterloo, fifty-four individuals with macular degeneration and fluent in English were invited to participate in the study. Nineteen participants passed the initial eligibility screen and agreed to participate. Six were ineligible due to beginning anti-VEGF treatments or having contraindications for tDCS. Five additional participants did not complete the study due to the COVID pandemic shutdown. Eight eligible participants completed the study, all with AMD (See Supp. Figure 1A).

At The Hong Kong Polytechnic University, fifty-three individuals with macular degeneration and fluent in reading Chinese characters were invited to participate in the study. Fifteen participants passed the initial eligibility screen and agreed to participate. Two participants did not complete the study due to complaints of inconvenient transportation to the laboratory. Thirteen eligible participants completed the study, two with JMD (one Stargardt disease and one Best disease) and eleven with AMD (See Supp. Figure 1B).

#### Visual stimuli

*Rapid Serial Visual Presentation (RSVP) Reading*

The RSVP test was created with custom-written software in python using the Psychopy library (Peirce et al., 2019). Using only their better eye, the other being occluded, participants fixated on a black cross against a bright background (~150 cd/m$^2$) at the beginning of each trial. A mask of "xxxxxxxxx" (English) or "xxx" (Chinese) was presented at the same exposure duration as each presented word or character at the start and end of each trial. Following the first mask, each word or character of a test sentence was presented separately in sequential order in the same position (See Supp. Video). Participants were asked to read out loud as many of the words in the sentence as possible. The viewing distance was 65 cm.

Participants read a variety of sentences in English or with Chinese characters. The English sentences were randomly selected from an existing pool of 2,627 sentences originally extracted from nine classic novels. All sentences had total lengths of between 40 and 80 characters, including spaces. All words were within the 5,000 most frequently written in English according to the British National corpus (Chung et al., 1998). The Chinese sentences were randomly selected from a pool of 605 sentences extracted from third-grade primary school textbooks. All sentences had a total length of 15 Chinese characters. No participant saw any individual sentence more than once.

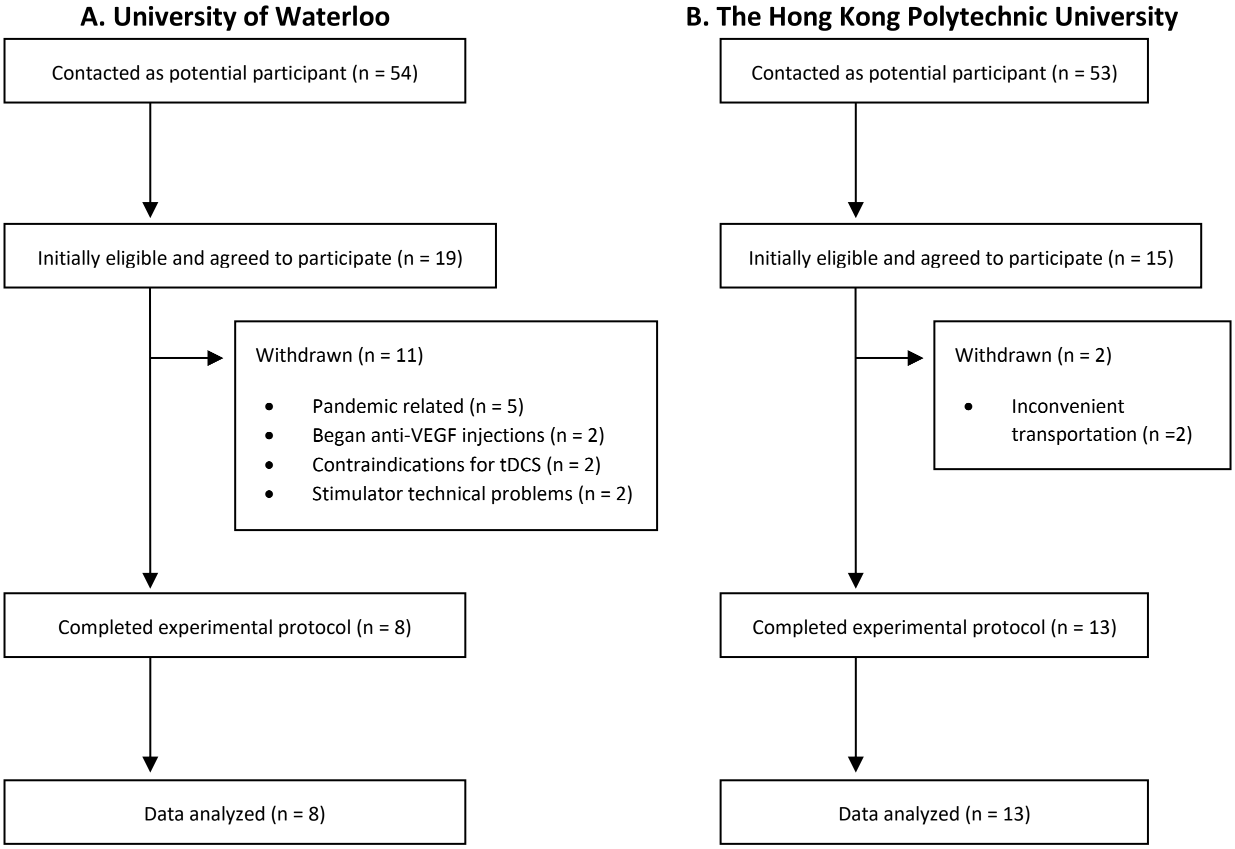


*Supp. Figure 1: Recruitment and participation flow diagram. A: University of Waterloo. B: The Hong Kong Polytechnic University.*

*Freiburg Visual Acuity Test (FrACT)*

Crowded and uncrowded visual acuity were assessed with the FrACT Landolt C test (Bach, 1996). Crowded tests employed a surrounding ring with 2 gap separation (twice the gap of the Landolt C). Participants verbally reported the orientation of the Landolt C, which randomly varied between the 4 cardinal and 4 oblique directions. The FrACT tests measured an acuity threshold using the Best PEST adaptive staircase procedure to target 56% accuracy. The viewing distance was 3 m.

### Experimental Procedure

*Clinical Screening*

Participants visited the laboratory once (Waterloo) or twice (Hong Kong) to complete screening and baseline measurements. An optometrist conducted a subjective trial frame refraction to identify the eye with the best corrected visual acuity. Distance VA was measured with 4m ETDRS original series charts 1 (right eye) and 2 (left eye) and scored by-letter. Participants were given an MNRead visual assessment for near reading visual acuity at 25cm corrected for that distance with a +4D addition in the trial frame. Reading speed in either English (Waterloo) or Chinese (Hong Kong) was scored using the manufacturer's instructions. The results of the MNRead test guided the initial stimulus parameters of the following RSVP baseline thresholding test (Cheong et al., 2007). Visual fields were examined with a modified tangent screen (Waterloo) or microperimetry (Hong Kong). In addition, participants underwent an eye examination to verify eligibility.

*Eye selection*

If the interocular difference in distance BCVA was ≥ 0.2 logMAR, then the eye with better VA was selected for testing during the experiment. Otherwise, the eye with the better MNREAD acuity was selected. If both eyes exhibited equal or nearly-equal MNREAD acuities, then the participant's preferred eye was selected, or the right eye if the participant had no preference.

*RSVP Baseline Thresholding*

During the initial baseline visit, participants performed a thresholding RSVP test. During this test, multiple print sizes and speeds were presented in order to find the 55% critical print size (CPS)– the smallest print size at which the participant could read at their maximum reading speed while achieving 55% accuracy. This CPS and associated maximum reading speed was exclusively used during the main treatment sessions. An accuracy of 55% was chosen to allow ample room for performance improvement.

The RSVP test was presented to the participant's selected eye, optically corrected for the 65cm viewing distance. Participants were presented with an initial RSVP print size and 5 different randomly interleaved reading speeds. The initial size tested was the CPS from the MNREAD test. The presentation times of each individual word or character on the screen (i.e. exposure duration) was determined by the maximum reading speed from the MNREAD. The initial five exposure durations were calculated as $ExposureDuration_{MNREAD\_equivalent} \times \left[\frac{1}{2.25}, \frac{1}{1.5}, 1, 1.5, 2.25\right]$, with slower reading speeds corresponding to longer exposure durations. One trial of each exposure duration was initially presented. If the reading performance elicited from these durations did not bound a range between 20%-80% correct, then the durations were adjusted. If the presented reading speeds elicited better (worse) performance than desired, then the $N$ slowest (fastest) speeds were replaced with $N$ new faster (slower) speeds, determined by dividing (multiplying) the adjacent exposure duration by 1.5. For example, if the original exposure duration array was $[A, B, C, D, E]$ and $N$ = 2 faster speeds were required, the exposure duration was updated to $[X, Z, A, B, C]$, where $Z = \frac{A}{1.5}$ and $X = \frac{Z}{1.5}$. $N$ was determined by the researcher's visual inspection of the accuracy plots (i.e. accuracy plotted as a function of exposure duration).

At least three trials of each duration from the final exposure duration array were completed, and a psychometric function was fitted to the data using a cumulative Gaussian curve. The procedure was then repeated on a new print size, either 0.16 logMAR larger than the current largest print size or 0.16 logMAR smaller than the current smallest print size, chosen by visual inspection based on the results of the previous print sizes. The initial 5 speeds were found by either multiplying or dividing the adjacent print size's exposure duration array by 1.3. In this way, slightly faster reading speeds would initially accompany each new larger print size, and slightly slower reading speeds would initially accompany each new smaller print size. Exposure durations were adjusted as described above if required (See Supp. Figure 2A-D). Additional print sizes were run until the session time limit had been reached (2 hours).

The print sizes (logMAR) and corresponding reading speeds eliciting 55% accuracy - converted to log words per minute - were fitted to a continuous bilinear piecewise function. This function contains one segment with a positive slope that increases with increasing print size up to a given point, after which a second segment with 0 slope extends for all larger print sizes. The intersection of both segments represents the CPS and maximum reading speed achieved by the individual participant. Because a point along the rising component of the curve is expected to be sensitive to improvements in both reading speed and CPS, the print size and associated reading speed selected for use during the RSVP treatment sessions corresponded to 0.2 logMAR below the fitted CPS (See Supp. Figure 2E-F). In one participant, the piecewise function did not fit the data any better than a standard linear fit (evaluated with the sum of squared residuals). Therefore, the average print size tested during thresholding, and the associated linearly-fitted reading speed, was selected for use during their RSVP treatment sessions.

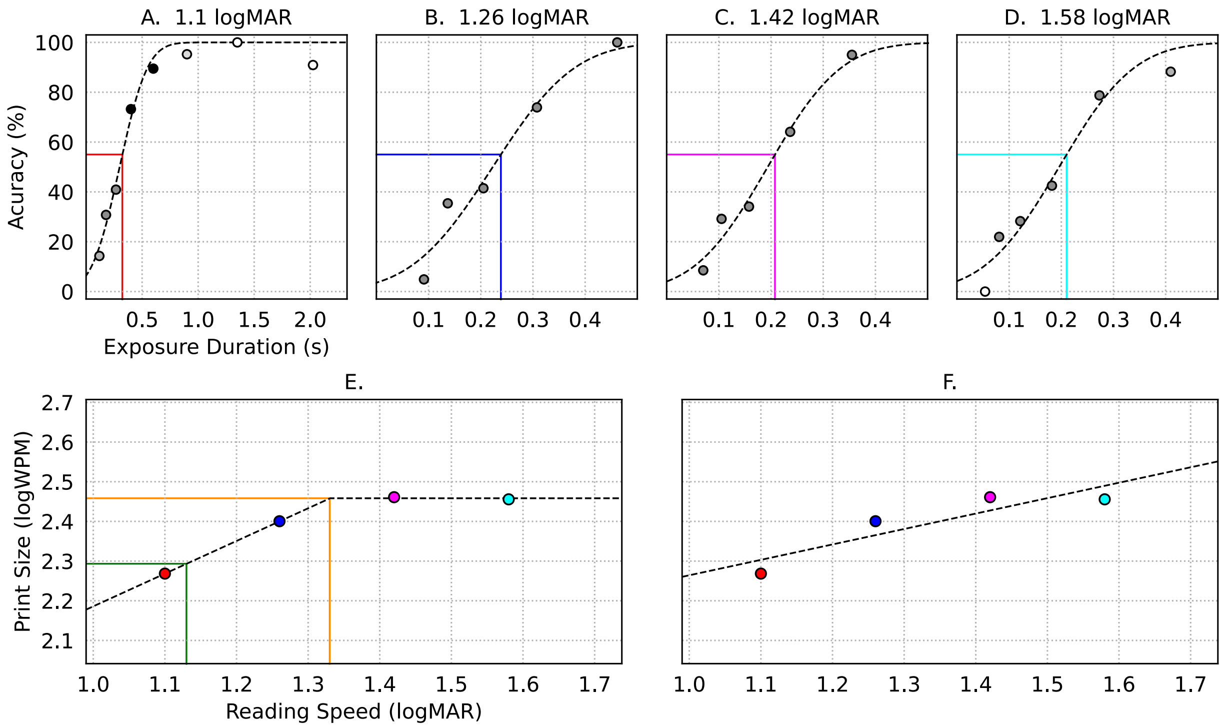


*Supp. Figure 2: Baseline RSVP curve fitting for an example participant. A-D. Accuracy for individual print size plotted against exposure duration (larger numbers indicate slower reading speed). Darker circles indicate conditions where more trials were run. White circles indicate conditions that were tested once before adjustment. The colored coordinates indicate the fitted points targeting 55% accuracy to be used during the continuous piecewise curve fit. E-F. Piecewise (E) and Linear (F) fits of reading speed against print size (in log-log scale). Colored circles correspond to the colored coordinates in panels A-D. The orange coordinate indicates the fitted critical print size (CPS) and maximum reading speed (MRS) targeting 55% accuracy. The green coordinate indicates the print size and reading speed used during the experimental treatment sessions which was 0.2 logMAR less than the CPS.*

*Treatment Sessions*

Participants completed two treatment sessions, one using active stimulation and the other using sham stimulation in a random order. Both experimenter and participant were blinded to the stimulation type. Treatment sessions were separated by at least 48 hours but no more than 7 days. First, a 15 sentence RSVP pre-test was presented at the reading speed and print size selected during RSVP thresholding to measure the primary outcome measure of reading performance accuracy. Afterward, crowded and uncrowded FrACT visual acuity thresholds in logMAR were measured as secondary outcome measures. A-tDCS was then started, and post-tests for all three outcome measures were run with identical settings to the pre-tests within the 20-minute stimulation time. This was repeated 5 minutes and then again 30 minutes after the stimulation was completed (See Supp. Figure 3).

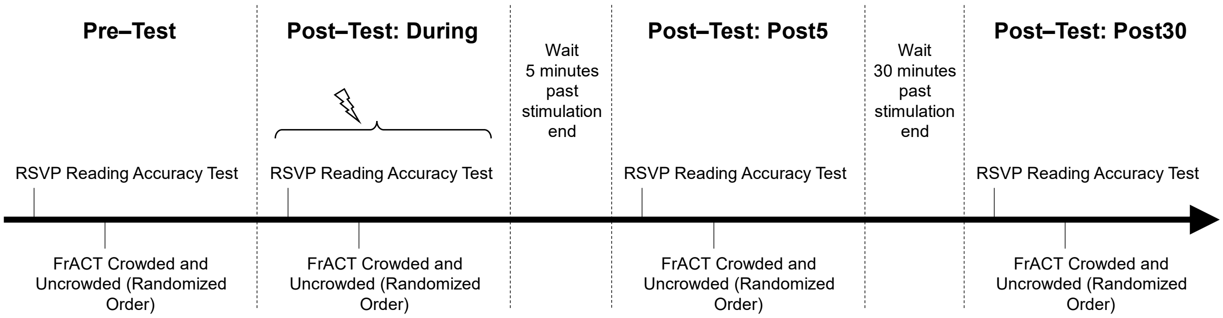


*Supp. Figure 3: Diagram of the treatment day procedure.*

**Data Analysis**

The data analysis was carried out according to a pre-specified data analysis plan (ClinicalTrials.gov ID: NCT04111068). All outcome measures were analysed with a 2 (Stimulation type: active, sham) × 3 (Test time: during stimulation, 5 minutes post stimulation, 30 minutes post stimulation) repeated measures ANOVA with language as a dichotomous covariate. All statistical analyses were run in JASP v. 0.16.1 (JASP Team, 2022). The effect of test time was calculated by subtracting pre-test performance from the given post-test performance (see main text). Therefore, a positive RSVP effect represents an improvement from baseline, while a negative crowded or uncrowded FrACT effect represents an improvement from baseline

In addition, follow-up ANOVAs with covariates of age, macular degeneration type, and baseline vision measures (distance VA, RSVP CPS, RSVP MRS) were run to determine if any significant effects were moderated by these covariates. To fairly compare across languages, all baseline vision covariates were created by rank ordering each participant separately within languages and performing a median split on the data, and corresponding halves between languages were combined. This created a balanced dichotomous separation between participants with better baseline vision measures from those with worse baseline measures across both testing sites.

**Supplementary Results**

*Secondary and Exploratory Analyses*

For crowded visual acuity, a significant main effect of test time was found, as well as a significant interaction between test time and language; $F(2,38) = 11.3$, $p < 0.001$, and $F(2,38) = 5.2$, $p = 0.008$, respectively. Importantly, a significant interaction between test time and stimulation type was also found, $F(2,38) = 3.7$, $p = 0.034$. Owing to this interaction effect, simple main effects of stimulation type were investigated, revealing a significant difference between the effect of active and sham stimulation at the "during" post-test ($During_{active} - PreTest_{active}$: 0.007 logMAR, SD: 0.096; $During_{sham} - PreTest_{sham}$: 0.040 logMAR, SD: 0.153), $F(1,19) = 6.5$, $p = 0.019$, but not at the other test times. When analyzing the languages separately, the main effect of test time was significant for the English-reading participants, $F(2,14) = 8.4$, $p = 0.004$. No significance was found for the participants reading Chinese characters. See Supp. Figure 4.

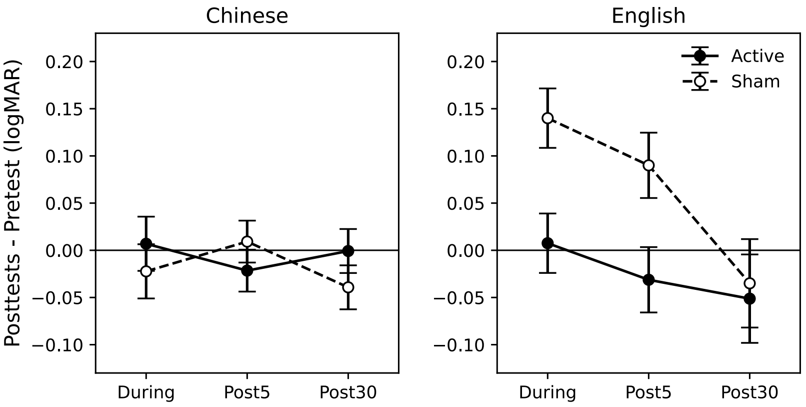


*Supp. Figure 4: The effect of a-tDCS on crowded visual acuity for Chinese readers (left plot) and English readers (right plot). The Freiburg Vision Test (FrACT) using the Landolt C optotype with a framing ring was used for both participant populations. Negative values indicate improved performance after stimulation. The Y-axis denotes the logMAR difference between the pre-test and each of the three post-tests. The error bars are ±1 SEM.*

When analyzing the uncrowded stimuli, no significant effects were found using language as a covariate. However, when using age as a covariate, a significant interaction between stimulation type and test time, and a significant three-way interaction was found, $F(2,38) = 3.3$, $p = 0.046$, and $F(2,38) = 3.3$, $p = 0.047$, respectively (See Supp. Figure 5A-B). These findings suggest that age moderated the effect of stimulation type when tested with uncrowded stimuli. In addition, when including either baseline vision measures of distance VA or RSVP CPS as covariates, a significant interaction was found between stimulation type and the covariate, RSVP CPS: $F(1,18) = 6.6$, $p = 0.019$, VA: $F(1,19) = 4.6$, $p = 0.046$, again suggesting a moderating effect of baseline visual ability on the effect of stimulation type (See Supp. Figure 5C-D).

Finally, a strength of crowding for visual acuity was calculated by subtracting each participant's crowded and uncrowded FrACT measurements, revealing only a significant effect of test time, $F(2,38) = 3.8$, $p = 0.030$, and an interaction between test time and language, $F(2,38) = 3.4$, $p = 0.044$, but no significant effects of stimulation.

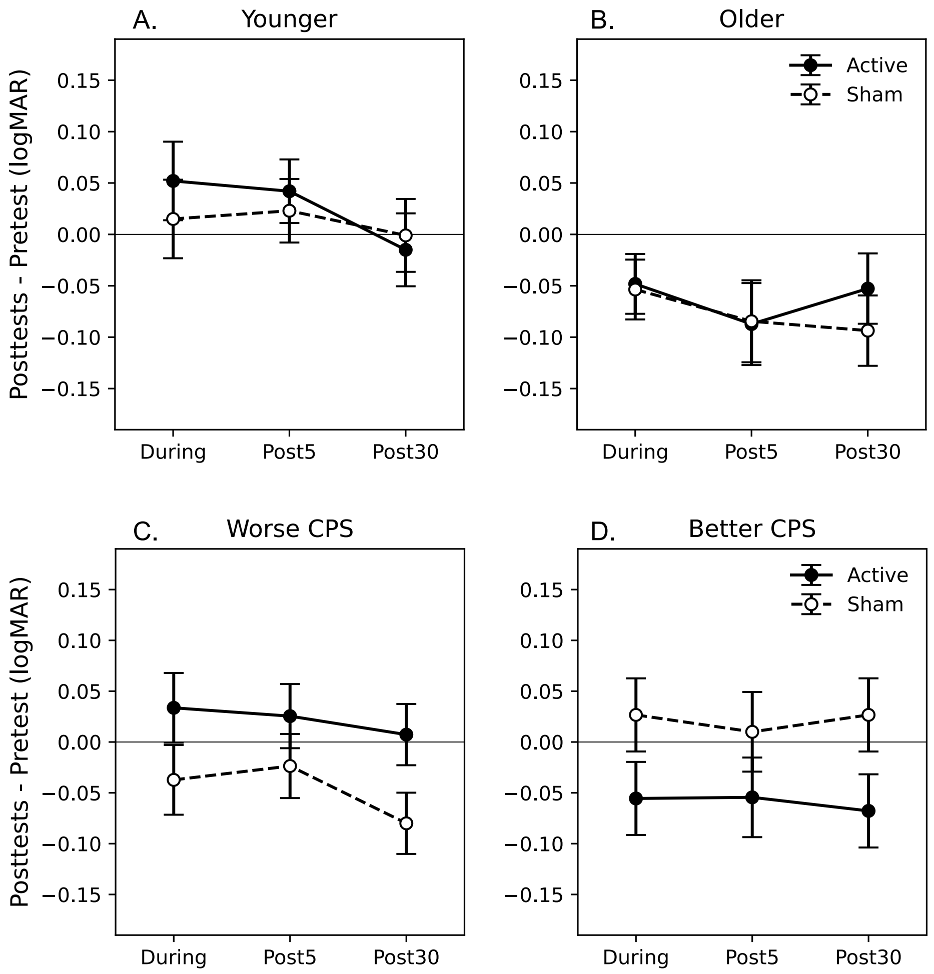


*Supp. Figure 5: The effect of a-tDCS on uncrowded visual acuity. Negative values indicate improved performance after stimulation. A-B. Participants were divided with a median split by age (median = 75.3). C-D. Participants were divided into worse and better critical print size groups. The participant split was similar when dividing along visual acuity, resulting in qualitatively similar results. All other conventions were the same as in Supp. Figure 4.*
